# Chloroquine and Hydroxychloroquine for the treatment of COVID-19: A Systematic Review and Meta-analysis

**DOI:** 10.1101/2020.07.04.20146381

**Authors:** Arunmozhimaran Elavarasi, Manya Prasad, Tulika Seth, Ranjit Kumar Sahoo, Karan Madan, Neeraj Nischal, Manish Soneja, Atul Sharma, Subir Kumar Maulik, Shalimar, Pramod Garg

## Abstract

**Background:** There is no effective therapy for COVID-19. Hydroxychloroquine (HCQ) and chloroquine (CQ) have been used for its treatment but their safety and efficacy remain uncertain.

**Objective:** We performed a systematic review to synthesize the available data on the efficacy and safety of CQ and HCQ for the treatment of COVID-19.

**Methods:** Two reviewers searched for published and pre-published relevant articles between December 2019 to 8th June 2020. The data from the selected studies were abstracted and analyzed for efficacy and safety outcomes. Critical appraisal of the evidence was done by Cochrane risk of bias tool and Newcastle Ottawa scale. The quality of evidence was graded as per the GRADE approach.

**Results:** We reviewed 12 observational and 3 randomized trials which included 10659 patients of whom 5713 received CQ/HCQ and 4966 received only standard of care. The efficacy of CQ/HCQ for COVID-19 was inconsistent across the studies. Meta-analysis of included studies revealed no significant reduction in mortality with HCQ use [RR 0.98 95% CI 0.66-1.46], time to fever resolution [mean difference −0.54 days (-1.19-011)] or clinical deterioration/development of ARDS with HCQ [RR 0.90 95% CI 0.47-1.71]. There was a higher risk of ECG abnormalities/arrhythmia with HCQ/CQ [RR 1.46 95% CI 1.04 to 2.06]. The quality of evidence was graded as very low for these outcomes.

**Author’s Conclusion:** The available evidence suggests that CQ or HCQ does not improve clinical outcomes in COVID-19. Well-designed randomized trials are required for assessing the efficacy and safety of HCQ and CQ for COVID-19..

## Background

A novel coronavirus-Severe Acute Respiratory Syndrome Coronavirus 2 (SARS-CoV-2) is responsible for the coronavirus disease 2019 (COVID-19). World Health Organization (WHO) declared COVID-19 as a global pandemic on 11^th^ March 2020. As of 8^th^ June, more than 7.1 million people have been infected, and 406959 have died due to COVID-19.[1] In the absence of any definitive therapy, repurposing of some commonly used antivirals and immunomodulatory drugs has been tried to treat COVID-19. Among such drugs, hydroxychloroquine (HCQ) was one of the first drugs to be tested for COVID-19 and has been recommended in national treatment guidelines in some countries such as India and the US [2]. Although the US FDA has not approved the drug for its clinical use, CDC (USA) has mentioned its use on the website as an approved drug lending support to such claims.[3] Clinical studies evaluating the efficacy of HCQ, have several limitations such as small sample size, heterogeneity, inconsistent results and early stoppage of trials and thus robust data are lacking with regard to its efficacy and safety. In view of the recent controversy, public anxiety and lack of effective therapy, it is therefore important to systematically review the literature, critically appraise it and present credible evidence, which might help treating clinicians, policy makers and patients make informed decisions. We performed a systematic review of studies that tested the efficacy of chloroquine (CQ) and HCQ for the SARS-CoV-2 infection/COVID-19 and did a meta-analysis of the relevant studies.

### Objective

This systematic review was carried out to answer the following research question:

In patients with COVID-19, is the use of CQ or HCQ effective and safe in reducing mortality and improving the clinical course, fever remission and virologic clearance as compared to no CQ/HCQ?

## Methods

Criteria for considering relevant studies for this review were as follows

### 1. Types of studies

We included studies on humans, which were randomized controlled trials (RCTs), prospective or retrospective case series or cohort studies with a control arm. We excluded case series without a control arm, case reports, review articles, viewpoints, experimental in vitro studies, editorials, and expert opinion. We included indexed studies from PubMed, Google scholar as well as non-indexed and pre-print articles from various pre-print servers because the latest information on COVID-19, a new disease, available on these pre-print servers might be valuable for the present review.

### 2. Types of Participants

We included human studies in which patients with confirmed COVID-19 of all ages and sexes were enrolled.

### 3. Types of Interventions

We sought studies in which patients were given HCQ or CQ in any dose, alone or combined with other drugs and had compared with patients in whom HCQ or chloroquine was not given.

### 4. Outcomes

For each study, we sought the following outcomes:

i. **Efficacy outcomes**
  ∘ **Clinical outcomes:** mortality, improvement in clinical course in terms of time to fever resolution, and development of acute respiratory distress syndrome (ARDS) or need for mechanical ventilation suggestive of worsening of disease.
  ∘ **Radiologic outcomes:** Improvement in findings on CT chest
  ∘ **Laboratory outcomes:** Virologic clearance as determined by RT-PCR test
ii. **Safety outcomes:** Adverse effects associated with HCQ/Chloroquine

### Search strategy and Identification of studies

Two authors independently searched the PubMed, Google Scholar and MedRxiv databases using the following search terms: “[(chloroquine OR hydroxychloroquine) AND (COVID-19 OR SARS-CoV-2)]” from 2000 to 8^th^ June 2020. Studies of all languages were included. No limits were applied to the search results except studies in humans. Hand searching of cross-references of original articles, reviews and pre-published articles was also performed to find additional relevant articles.

### Data collection and analysis

The citations were retrieved into a reference management software (Zotero version 5.0.85). Duplicate citations were removed. All the remaining studies were reviewed by going through their title and abstract to select the studies meeting our inclusion criteria mentioned above. Data on outcomes were extracted by one reviewer (AE) and cross-checked by another reviewer (Sh). The risk of bias was assessed using the Cochrane Risk of Bias Tool for RCTs[4] and the Newcastle Ottawa Scale for Cohort studies.[5] Meta-analysis of RCTs was done for the outcomes virologic clearance and time to fever resolution. Meta-analyses of the other studies were done for the following outcomes:

1. Mortality,
2. Worsening of clinical condition in the form of development of ARDS or need for mechanical ventilation,
3. Virologic clearance
4. ECG abnormalities and de novo ventricular arrhythmias.

We calculated relative treatment effects using Mantel-Haenszel random-effects model, expressed as RRs with 95% (CIs) for the outcomes: mortality, worsening of pneumonia/ARDS/mechanical ventilation, and virologic clearance. For the outcome ECG abnormalities/de novo ventricular arrhythmia, odds ratio(OR) with 95% CI was calculated using the generic inverse variance method. Adjusted odds ratio was used whenever available and unadjusted odds ratio in other circumstances. The standard error was calculated from the 95% confidence intervals. In the case of continuous variables, the mean difference for each study was plotted, and the summary statistic was calculated using the inverse variance method. All the analyses were performed using RevMan 5.3.

The certainty in the evidence was graded using the GRADE methodology (Grading of Recommendations, Assessment, Development and Evaluations).[6]

## Results

### Description of studies

The PubMed search yielded 579 articles. MedRxiv search yielded 291 articles and Google scholar search yielded 1360 articles. The Cohen’s Kappa for inter-rater agreement between the 2 reviewers was 0.8. Hand searching of relevant articles yielded one article. After excluding articles that did not fulfill the inclusion criteria and duplicate citations, we had 20 articles to be examined in detail.

Out of these 20, one study was excluded as it compared two doses of HCQ, the comparison was with historical data, and complete information was missing.[7] Another article was excluded as it had incomplete outcome data.[8] Two studies were excluded as the authors had withdrawn the papers.[9,10] Another study was excluded as the exposure data and co-interventions were incompletely reported.[11] Thus, 15 articles [12–26] were included for the final review and synthesis of evidence, and assessment of the risk of bias. PRISMA (Preferred Reporting Items for Systematic reviews and Meta-Analyses) flow chart has been provided (Figure 1)

**Figure 1:**
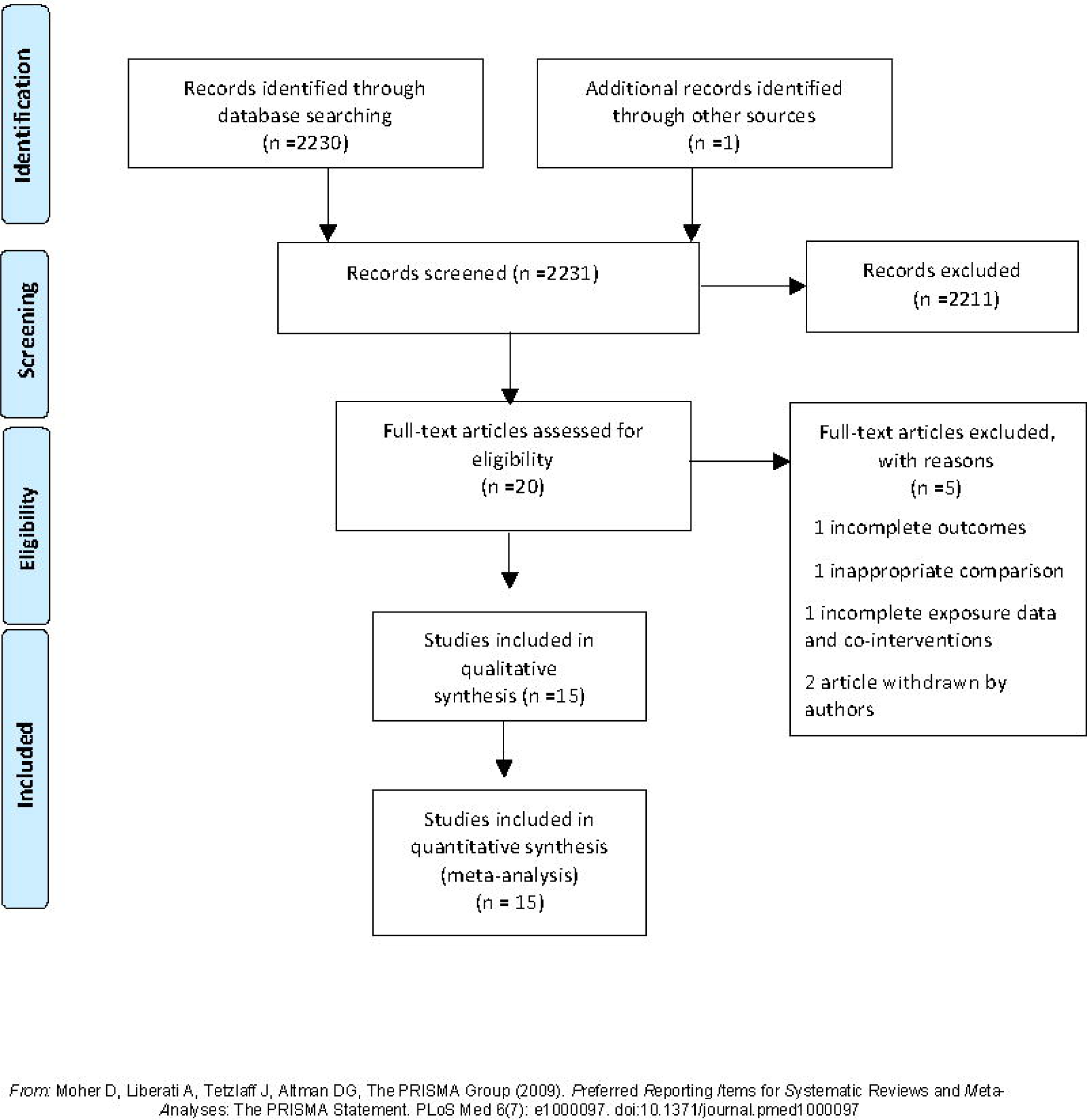
PRISMA flow diagram for study selection

### Studies included

The 15 studies that were selected had included 10659 patients. Of these, 12 were cohort studies which had a control group and 3 RCTs. The details about the study design, participants, interventions and outcomes are described in Tables 1-3.

**Table 1:** Summary Results of the included cohort studies with mortality as outcome.

|  |  |  |  |  |  |  |  |  |  |
| --- | --- | --- | --- | --- | --- | --- | --- | --- | --- |
| S. No. | Author | Type of study | Total Patients (n) Age median (range, years) Co-morbidity | Controls (no HCQ) (n) | Severity of illness | Intervention (HCQ); number of patients, Dose, duration | Concomitant medicines | Outcome | Overall benefit |
| 1 | Bo Yu et al. | Retrospective Cohort | 568 patients Median age 68 years (Range 57-76) | 520 | Critically ill COVID-19 patients with severe ARDS with PaO<sub>2</sub>/FiO<sub>2</sub> <300 on mechanical ventilation | n=48 HCQ 200 mg BD x 7-10 days | received antivirals and supportive care including IVIg, antibiotics and interferons | Mortality 18.8% (9/48) in HCQ group and 45.8% (238/520) in non-HCQ group (p<0.001) | Yes |
| 2 | Geleris et al. | Prospective Cohort | 1376 Patients with COVID-19 who were hospitalized | 565 | not intubated for at least 24 hours after starting of HCQ | n=811 HCQ 600 mg on day 1 followed by 400 mg for 4 days; | Azithromycin 500 mg on day 1 followed by 250 mg for 4 days | Respiratory failure needing intubation or death in 252/811 (31%) of HCQ vs 84/565 (14.8%) of non HCQ group (HR 1.04, 95% CI 0.82 to 1.32)* | No |
| 3 | Ip et al. | Retrospective cohort | hospitalized COVID-19 n=2512 | 598 | 37% had O<sub>2</sub> saturation <94% | N=1914 HCQ 800 mg on day 1, and 400 mg on day 2-5 | 77% received azithromycin | Mortality 432/1914 (22.5%) in HCQ arm and 115/598 (19.2%) in non HCQ arm. Propensity-score adjusted hazard ratios, any use of hydroxychloroquine during the hospitalization (HR, 0.99; 95% CI, 0.80-1.22). Arrhythmias 5% in HCQ and 4% in non HCQ arm | No |
| 4 | Magagnoli et al, | Retrospective cohort | 368 Hospitalized patients | 158 | 136/368 (36.9%) had O<sub>2</sub> saturation <94% | n=210 HCQ alone, n=97 HCQ+AZ n=113 | Standard of care | Mortality in HC, HC+AZ, and no HC groups were 27.8%, 22.1%, 11.4%, respectively. Risk of death higher in HCQ group (HR 2.61; 95% CI, 1.10 to 6.17; P=0.03) but not in HCQ+AZ (HR 1.14; 95% CI 0.56—2.32; p=0.72) Need for ventilation similar in HCQ vs no HCQ | Higher deaths in HCQ group |
| 5 | Mahevas et al | Retrospective Cohort | 181 Adults, median age 60 | 97 | oxygen requirement | n=84 HCQ 600 | Standard of care | 20.2% in HCQ group transferred to the ICU or died within 7 days vs | No |
|  |  |  | years (IQR 52-68 years); comorbidities lesser in HCQ group |  | >2L/min | mg/day in first 48 hours of hospitalization |  | 22.1% in the no-HCQ group (RR 0.91, 95% CI 0.47–1.80); 27.4% in HCQ and in non HCQ group 24.1%, respectively, developed ARDS within 7 days (RR 1.14, 95% CI 0.65–2.00) |  |
| 6 | Membri llo et al. | Prospective cohort | n=168 Hospitalized Age: 18-85 years | 43 | All patients irrespective of severity | N=123 HCQ Loading dose of 800 mg followed by 400 mg per day of HCQ | Standard of care which included antivirals, immunomodulation including steroids/tocilizumab | Mortality 27/123 (22%) in the HCQ arm vs. 21/43 (48.8%) in the standard of care arm [p=0.002] | Yes |
| 7 | Rosenberg et al. | Retrospective cohort | n=1438 median age 63 years, Comorbidities in 60%, commonest hypertension | 221 | All hospitalized patients, severity not mentioned | HCQ alone or with azithromycin n=1006 HCQ+AZ n=735 | Standard of care | Mortality HCQ alone 54/271 (19.9%), vs. HCQ+AZ 189/735 (25.7%) vs. 243/1006 in HCQ vs. 28/211 (12.7%). Overall no difference in mortality for HCQ+AZ (HR, 1.35 [95% CI, 0.76-2.40]), HCQ alone (HR, 1.08 [95% CI, 0.63-1.85]).<br>In regression models of abnormal ECG findings, no significant differences between the groups receiving neither drug vs. HCQ+AZ and HCQ alone | No |
| 8 | Singh et al. | Retrospective cohort | n=1125 (N=910 for propensity matched analysis); Mean age 61.45 years | N=2247; (N=910 for propensity matched analysis) | Severity not mentioned | HCQ dose and duration not mentioned; 799 received azithromycin | controls received standard of care/Azithromycin/corticosteroids and no other antivirals or biologics | 30 day mortality 104/910 (11.8%) in HCQ vs 109/910 (11.9%) in Standard of care arm (RR 0.95; 95% CI 0.74,1.23) (Propensity score matched sample) Mech. ventilation 46/910 in HCQ vs 57/910 in standard of care | No |

\*This study evaluated death or intubation as a composite outcome.
CQ: chloroquine; HCQ: hydroxychloroquine; IVIg: intravenous immunoglobulin; Az: azithromycin

**Table 2:** Summary Results of the included cohort studies with virologic clearance as the outcome.

|  |  |  |  |  |  |  |  |  |  |
| --- | --- | --- | --- | --- | --- | --- | --- | --- | --- |
| S. No. | Author | Type of study | Patients (n)<br>Age median (range) years<br>Co-morbidity | Controls (no HCQ) (n) | Severity of illness | Intervention Number, dose | Concomitant medicines | Outcome | Overall benefit |
| 1 | Gautret et al | Cohort study with controls from other hospital | 42 Hospitalized patients aged >12 years with SARS-CoV-2 positive | 16 | All stages of COVID | N=26<br>HCQ 200 mgTDSx10 days | Azithromycin | Virologic clearance at day 6 was 14/20 in the HCQ group vs 2/16 in the non HCQ group | Yes |
| 2 | Jihad Mallat et al | Retrospective cohort study | 34 Adult patients, 10 had comorbidities, all of them survived | 13 | 6 required oxygen, none required high flow oxygen or ICU care | n=21<br>HCQ 400 mg BD first day followed by 400 mg OD x 10 days | Standard of care | On day 14, 47.8% (14/23) patients tested negative in the HCQ group compared to 90.9% (10/11) patients who did not receive HCQ (p=0.016) | No |
| 3 | Huang et al. | Prospective cohort | 233 adult patients with COVID-19 with median age 43 (33-55) years | 176 | Critical cases excluded | n=197<br>Chloroquine 500 mg OD (low dose) or BD (high dose) | Standard of care | By day 10, 91.4% of CQ and 57% of non CQ had negative swab and day 14 95.9% of CQ vs 79.6% non CQ had negative swab | Yes |

**Table 3:** Summary Results of the 3 RCTs on HCQ in COVID-19.

| Table 3: Summary Results of the 3 RCTs on HCQ in COVID-19 |  |  |  |  |  |  |  |  |  |
| --- | --- | --- | --- | --- | --- | --- | --- | --- | --- |
| S. No. | Author | Type of study | Patients (n)<br>Age<br>Co-morbidity | Controls (no HCQ) (n) | Severity of illness | Intervention Number, dose | Concomitant medicines | Outcome | Overall benefit |
| 1 | Chen Jun et al. | RCT | 30 treatment Naive COVID-19 patients | 15 | Not available | N=15<br>HCQ 500 mg/d x 5 days | NA | On day 7, throat swab was negative in 13 (86.7%) cases in the HCQ group and 14 (93.3%) cases in the control group ($P > 0.05$).<br>Median time to fever remission HCQ group was 1 (0-2) vs control group 1 (0-3).<br>Radiological progression was shown on CT images in 5 cases (33.3%) of the HCQ group and 7 cases (46.7%) of the control group | No |
| 2 | Wei Tang et al. | RCT | N=150<br>Adult hospitalized patients positive for SARS CoV-2 | 75 | All patients | n=75<br>HCQ 1200 mg/d x 3 days followed by 800 mg x 2-3 weeks | Standard of care | 28-day negative conversion rate was not different between HCQ and control group (85.4% versus 81.3%, $P=0.34$) | No |
| 3 | Chen et al. | RCT | N=62<br>Adult patients | 31 | Mild COVID-19 | n=31<br>HCQ 400 mg/day | Standard of care | Compared with the controls [3.2 (1.3) days], the body temperature recovery time was significantly shortened with HCQ [2.2 (0.4) days]<br>CT chest improved on day 6 in HCQ group (80.6%, 25 of 31) compared with the control group (54.8%, 17 of 31) | Yes |

### Risk of Bias in included studies

Assessment of bias for the 3 RCTs and 12 cohort studies revealed a significant risk of bias as assessed against various quality parameters. Most of the comparative studies were of poor methodologic quality and were subject to high risk of bias owing to the non-randomized study design and the lack of placebo control. Even the RCTs were unblinded and subject to risk of assessment bias. The details are shown in Figure 2 and Table 4.

**Table 4:**
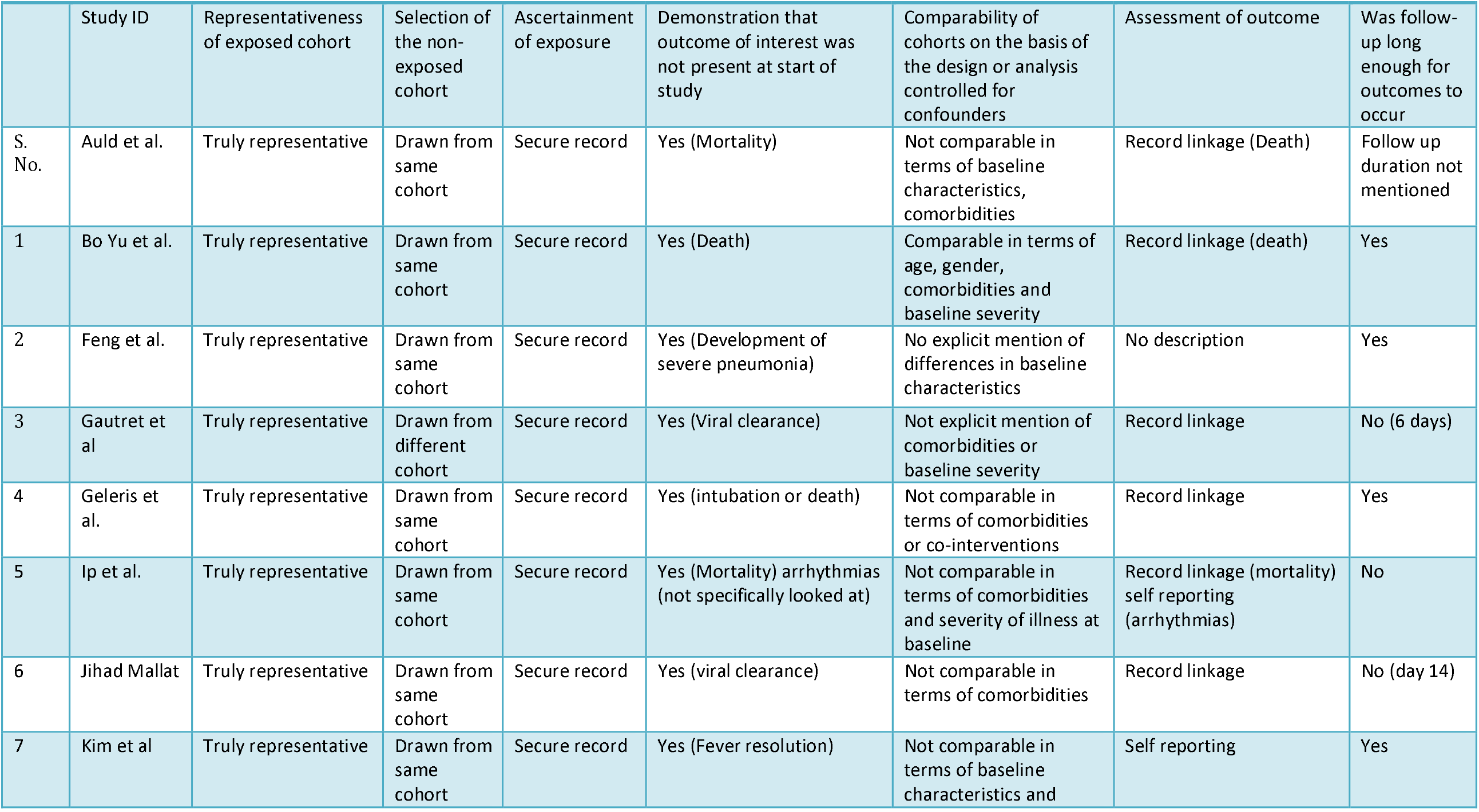
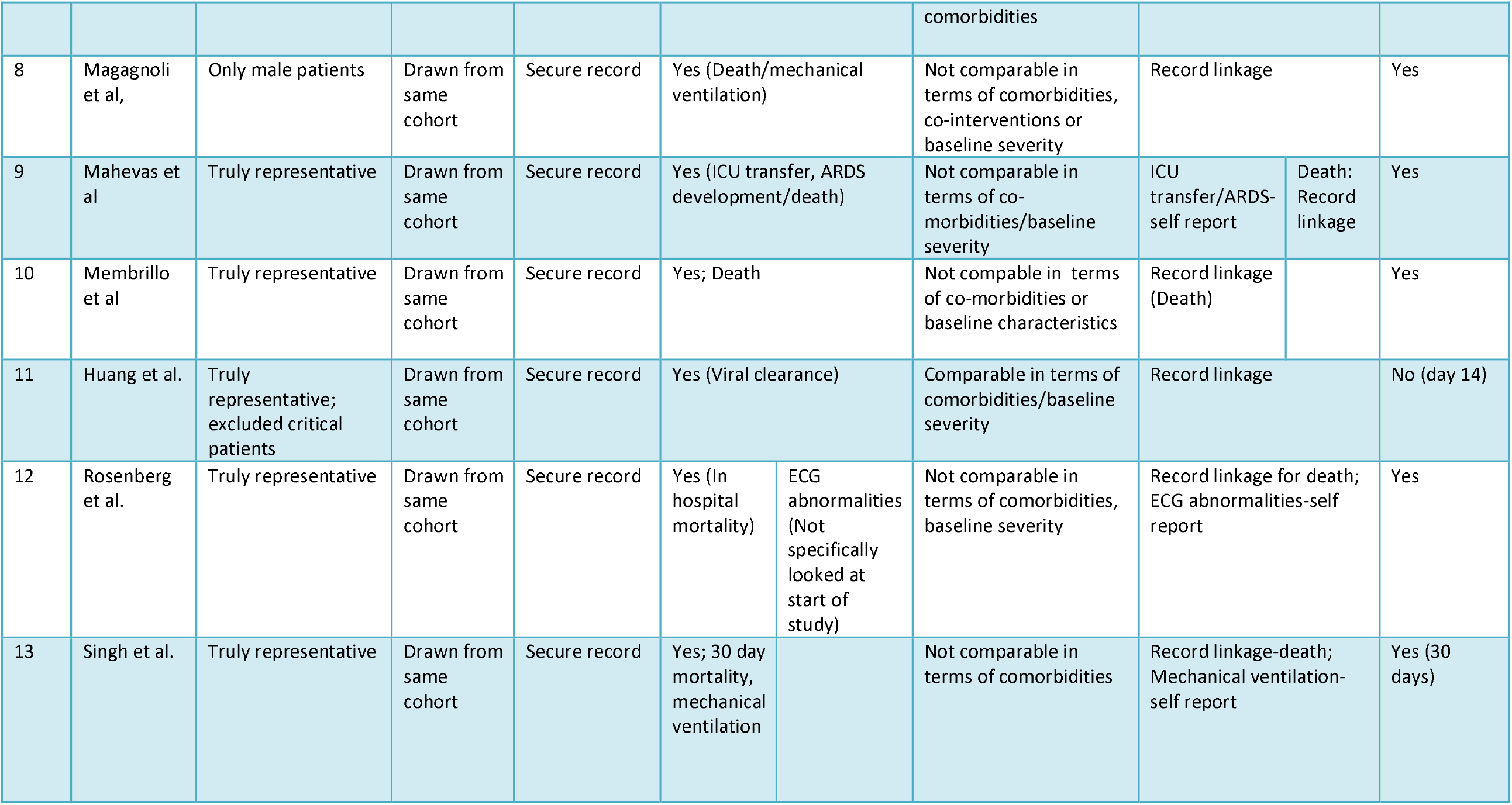
Newcastle-Ottawa Risk of bias assessment for cohort studies.

**Figure 2:**
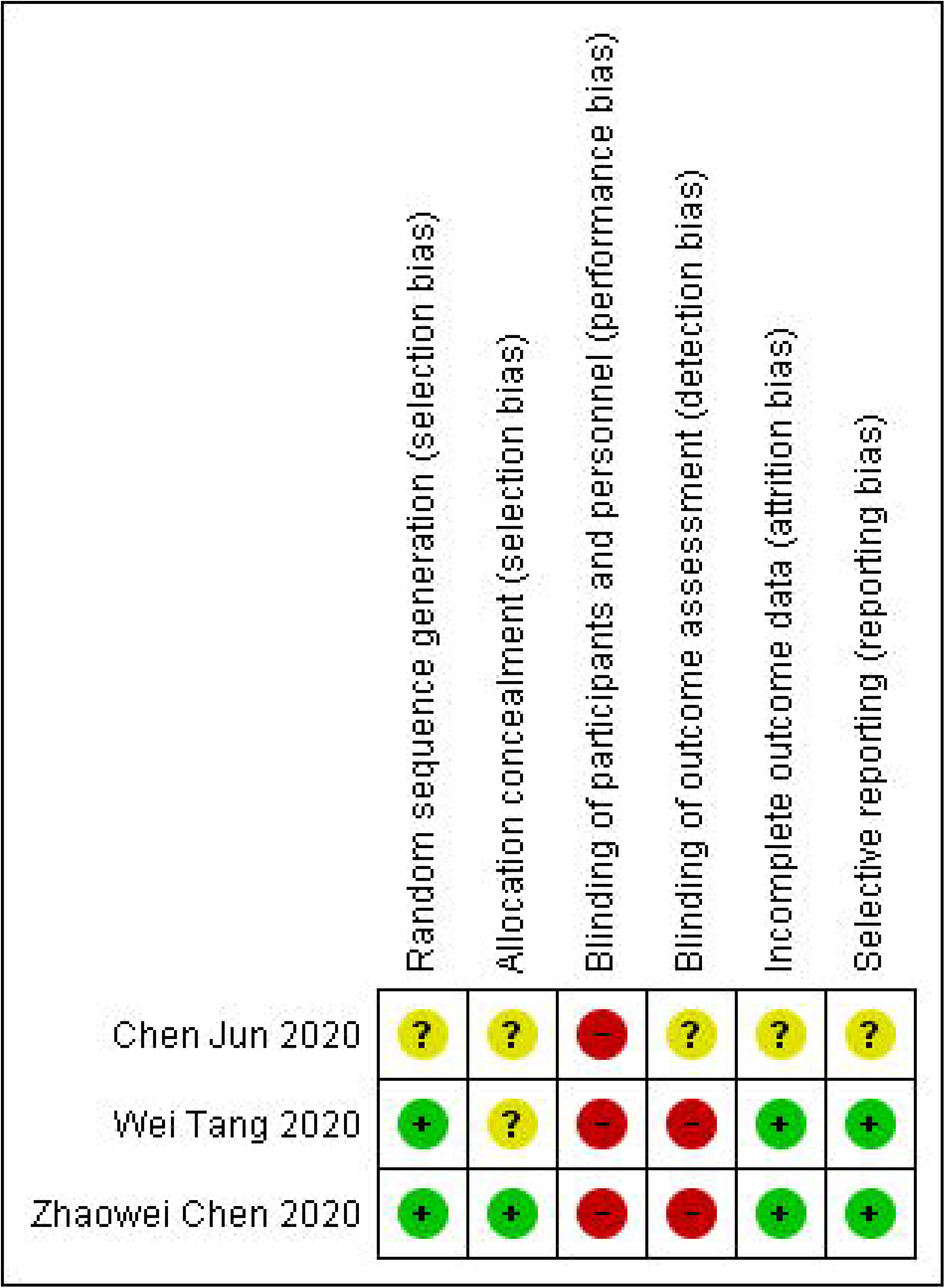
Risk of bias assessment for included RCTs

### Effects of CQ and HCQ on COVID-19

Of the 15 studies, 2 studies reported the use of CQ and 13 reported the use of HCQ. Of the total 10659 patients included in 15 studies, 5713 received CQ or HCQ along with standard of care and 4946 received only standard of care (no CQ/HCQ). Each study reported only a few of the outcomes of interest.

### Efficacy Outcomes

#### Mortality

7 cohort studies reported mortality. Two of them showed a reduction in mortality, 3 did not show any benefit and 2 showed higher mortality with the use of HCQ compared to controls(Table 1). However on after adjusting for baseline severity and comorbidities, one of the cohort studies did not show any difference in mortality with the use of HCQ.[21]

#### Clinical Course

One RCT and 5 cohort studies reported the effect of CQ/HCQ on the clinical course of the disease with either improvement or worsening in the form of development of ARDS or requirement for mechanical ventilation. One cohort study reported higher disease progression, one RCT and 4 cohort studies showed no difference in clinical course between CQ/HCQ and standard of care arm.

#### Time to fever remission

Two RCTs and one cohort reported this outcome. One of the RCTs reported a shorter time to fever remission with the use of HCQ while the other RCT and a cohort study did not.

#### Virologic clearance

Three cohort studies and 2 RCTs reported on virologic clearance. Two cohort studies found faster clearance of viral RNA in the HCQ group as compared to the standard of care group at study endpoints. But both the RCTs did not show any benefit in terms of virologic clearance (Table 2).

#### Safety outcomes

Two cohort studies reported ECG abnormalities and one of them reported a higher occurrence of ECG abnormalities in patients receiving HCQ with or without azithromycin. However, after adjusting for age, gender, comorbidities and baseline severity, there was no significant increase in odds of ECG abnormality. The other cohort study reported no difference in cardiac arrhythmias.

#### Meta-analysis

Meta-analysis of 7 studies showed no significant reduction in mortality with HCQ use [RR 0.98 95% CI 0.66-1.46] (Figure 3). There was no significant difference with regard to clinical deterioration/development of ARDS/need for mechanical ventilation between the HCQ and standard of care [RR 0.90 95% CI 0.47-1.71] (Figure 4). There was no statistically significant difference in virologic clearance between HCQ and placebo in the meta-analysis of 2 RCTs and 3 cohort studies [RR 1.03, 95% CI 0.83-1.28] (Figure 5). The time to fever remission was reported in 2 RCTs and one cohort study; meta-analysis showed reduced time to fever remission in the HCQ arm [mean difference −0.54 days, 95% CI -1.19 to 0.11], which did not attain statistical significance. (Figure 6)

**Figure 3:**
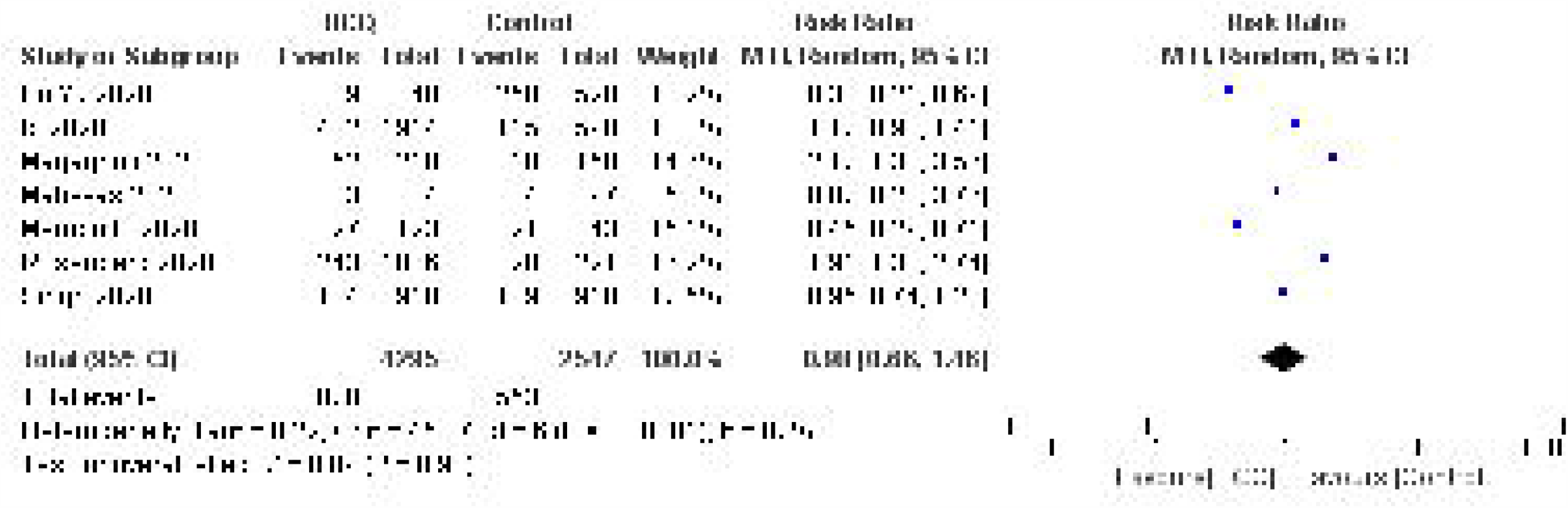
Effect of HCQ on mortality in patients with COVID-19

**Figure 4:**
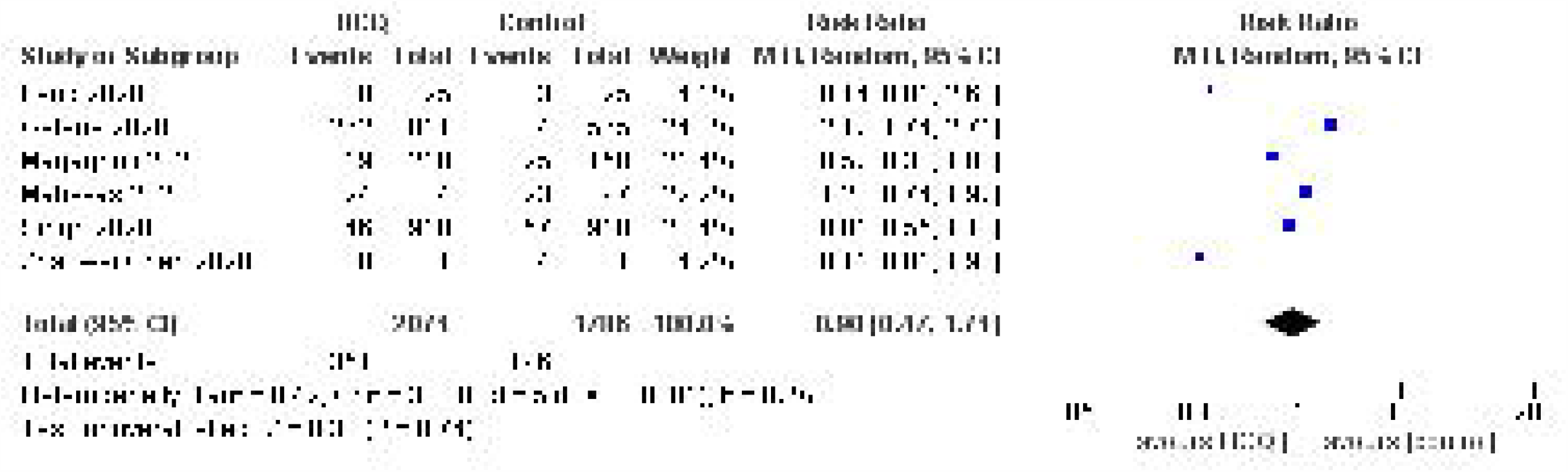
Effect of HCQ on clinical deterioration/need for mechanical ventilation in patients with COVID-19

**Figure 5:**
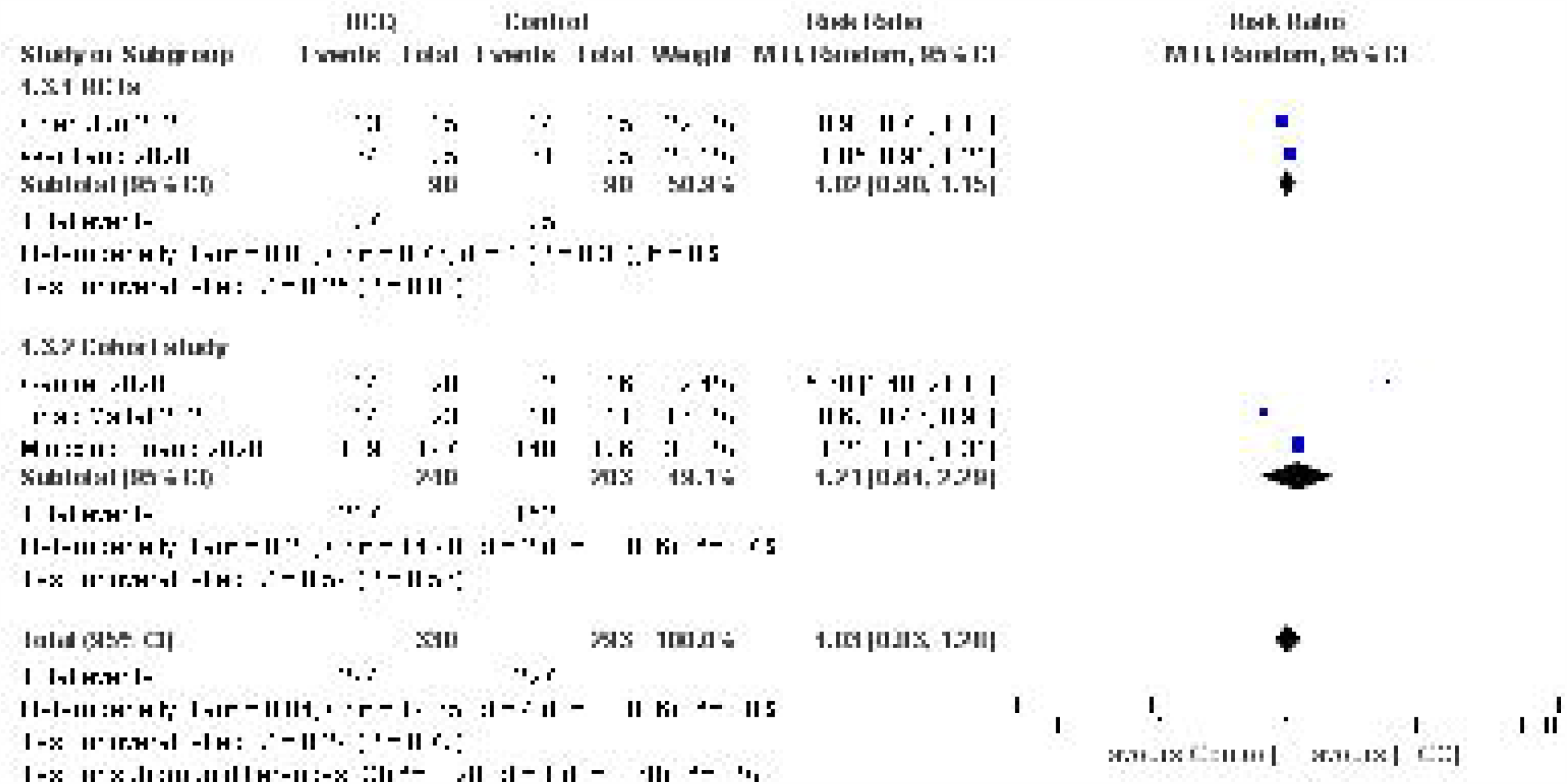
Effect of HCQ on virological clearance in patients with COVID-19

**Figure 6:**
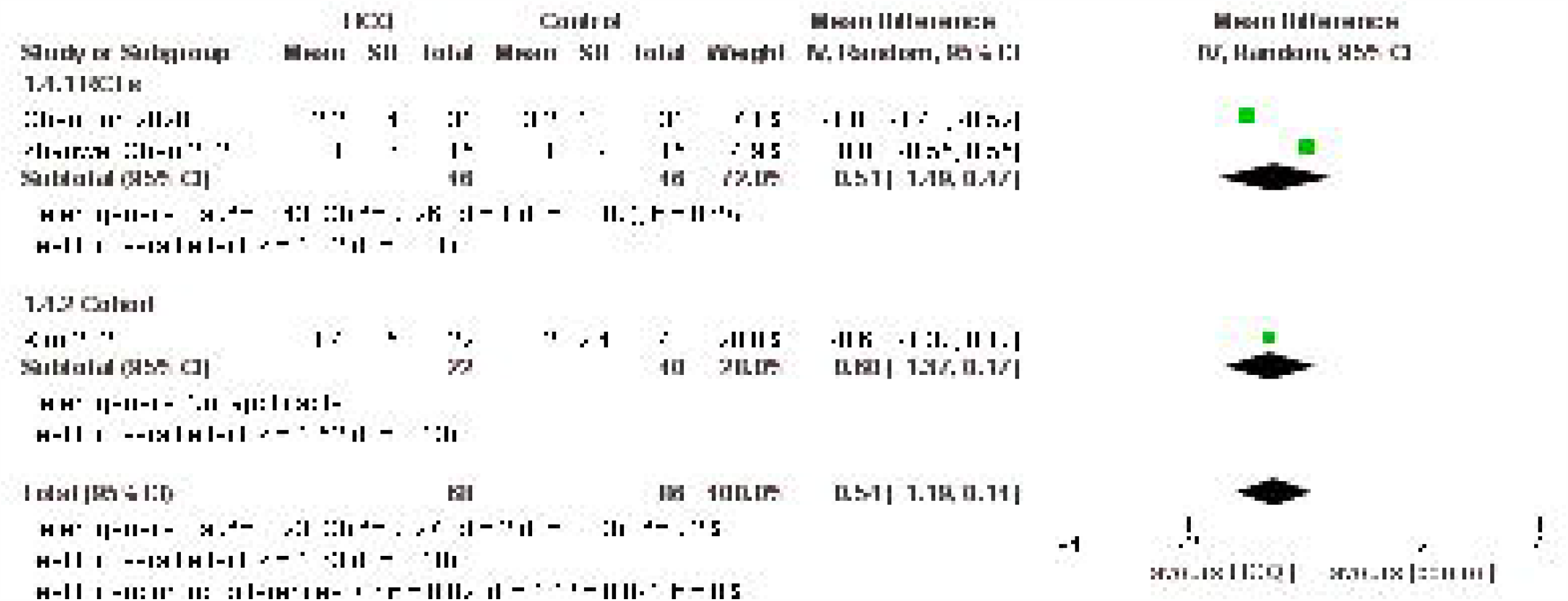
Effect of HCQ on time to fever remission in patients with COVID-19

Meta-analysis of two cohort studies suggested an increased risk of ECG abnormalities/cardiac arrhythmias in the HCQ arm as compared to standard of care and this was statistically significant (RR 1.46 [95% CI 1.04-2.06]. (Figure 7)

**Figure 7:**
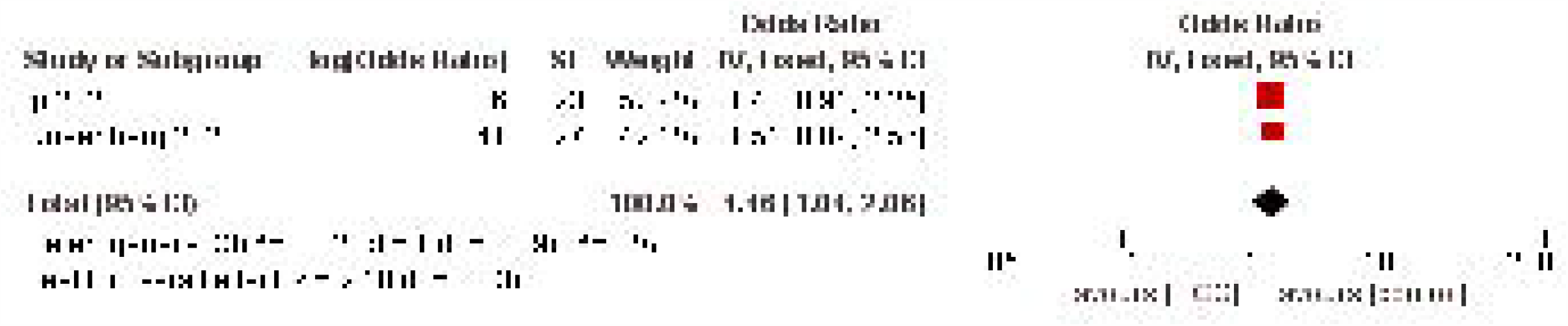
Effect of HCQ on ECG abnormalities in patients with COVID-19

#### Critical Appraisal

We included all clinical studies in humans in which HCQ- or CQ-arm was compared with no HCQ- or CQ-arm. We found significant heterogeneity in the inclusion criteria of the studies in which patients ranged from asymptomatic to severe and critically ill COVID-19. The other causes of heterogeneity were the dosage of HCQ, the use of other supportive care interventions including corticosteroids, antiviral drugs, tocilizumab, and IVIG. The stage of disease at which the drug was administered and host factors such as age, comorbid conditions were also different between the various studies. Similarly, virologic clearance was checked at various time points from day 6 to day 28. There was a significant risk of bias due to the study design being non randomized in 12 of the 15 included studies. The RCTs were not blinded and fraught with a risk of assessment bias. The reason for giving HCQ/CQ in some patients and not giving in others was not explicitly mentioned in any of the cohort studies. The Cochrane risk of bias tool was used to critically appraise the RCTs and Newcastle Ottawa Scale to assess the risk of bias in cohort studies and the risk is summarized in Figure 2 and Table 4.

#### Quality of Evidence

The evidence was judged to be of very low quality for the outcomes mortality, clinical deterioration/ARDS/need for mechanical ventilation, virologic clearance (cohort studies), time to fever resolution (cohort studies) and ECG abnormalities. For the outcomes virologic cure (RCTs) and time to fever remission (RCTs) it was judged to be low quality (Table 5).

**Table 5:** Hydroxychloroquine compared to placebo for COVID-19.

| Outcomes | No of participants (studies)<br>Follow up | Certainty of the evidence (GRADE) | Relative effect (95% CI) | Anticipated absolute effects |  |
| --- | --- | --- | --- | --- | --- |
|  |  |  |  | Risk with placebo | Risk difference with Hydroxychloroquine |
| **Death** | 6904 (8 observational studies) | ⊕○○○<br>VERY LOW | **RR 0.98**<br>(0.66 to 1.46) | 214 per 1,000 | 4 fewer per 1,000<br>(73 fewer to 98 more) |
| **Worsening of disease, Need for ventilation or development of ARDS** | 3857 (6 observational studies) | ⊕○○○<br>VERY LOW | **RR 0.90**<br>(0.47 to 1.71) | 110 per 1,000 | 11 fewer per 1,000<br>(58 fewer to 78 more) |
| **Virologic clearance - RCTs** | 180 (2 RCTs) | ⊕⊕○○<br>LOW | **RR 1.02**<br>(0.90 to 1.15) | 833 per 1,000 | 17 more per 1,000<br>(83 fewer to 125 more) |
| **Virologic clearance - Cohort study** | 443 (3 observational studies) | ⊕○○○<br>VERY LOW | **RR 1.21**<br>(0.64 to 2.29) | 749 per 1,000 | 157 more per 1,000<br>(270 fewer to 966 more) |
| **Time to fever remission - RCTs** | 92 (2 RCTs) | ⊕⊕○○<br>LOW | - | The mean time to fever remission - RCTs was 0 | MD 0.51 lower<br>(1.49 lower to 0.47 higher) |
| **Time to fever remission - Cohort** | 62 (1 observational study) | ⊕○○○<br>VERY LOW | - | The mean time to fever remission - Cohort was 0 | MD 0.6 lower<br>(1.37 lower to 0.17 higher) |
| **ECG abnormalities** | 3534 (2 observational studies) | ⊕○○○<br>VERY LOW | **RR 1.46**<br>(1.04 to 2.06) |  |  |

\*The risk in the intervention group (and its 95% confidence interval) is based on the assumed risk in the comparison group and the relative effect of the intervention (and its 95% CI).
CI: **Confidence interval**; RR: **Risk ratio**; MD: **Mean difference**
GRADE Working Group grades of evidence
High certainty: **We are very confident that the true effect lies close to that of the estimate of the effect**
Moderate certainty: **We are moderately confident in the effect estimate: The true effect is likely to be close to the estimate of the effect, but there is a possibility that it is substantially different**
Low certainty: **Our confidence in the effect estimate is limited: The true effect may be substantially different from the estimate of the effect**
Very low certainty: **We have very little confidence in the effect estimate: The true effect is likely to be substantially different from the estimate of effect**

## Discussion

Due to the sheer magnitude of the COVID-19 pandemic and lack of effective therapy, there is a race to find therapies that would improve the clinical outcomes of the patients. Amongst the various medications tried, HCQ has received maximum attention, partly due to the media coverage. A few initial in-vitro studies as well as a proof of concept study by Guatret *et al*. had shown some benefit [15]. Subsequently, many institutional protocols in countries like China, France, US and India[2] mandated the use of HCQ in the management of all patients with COVID-19. This has led to several observational studies being reported in rapid succession, which were of poor methodologic quality and most did not report outcomes of clinical interest uniformly. There were 3 RCTs but neither of them reported outcomes in terms of mortality, need for mechanical ventilation or ECG abnormalities. The initial observational studies and RCTs had many limitations such as small sample size, heterogenous patient population, variable endpoints, variable dosing of the drug, and no control for confounders.

Serious adverse drug reactions with the use of CQ and HCQ though uncommon have been reported. These include cardiac toxicity in the form of cardiomyopathy and prolonged QTc interval,[27] and hemolysis in patients with underlying G6PD deficiency. Moreover, their therapeutic index is narrow.[28] The NIH panel has recommended against using high-dose chloroquine (600 mg twice daily for 10 days) and a combination of hydroxychloroquine plus azithromycin because of the potential for toxicities.[29] The most recent and largest study which reported a higher mortality was subsequently retracted due to multiple reasons chiefly lack of access to data for independent review held by a private company.[30] The investigators of a multicenter trial, the RECOVERY trial, being conducted in the UK have announced negative results but the full report is still to be published.

Our results are at odds with those of a recent meta-analysis from France which has shown that HCQ results in significant improvements in various clinical parameters including mortality.[31]. However, this meta-analysis did not use the standard methodology for a meta-analysis and did not do a proper risk of bias assessment for the included studies. They also included a study in their meta-analysis which has now been retracted. The quality of evidence was also not graded. At this time, it is important to present credible evidence due to recent scientific controversy, political discourse, and heightened public anxiety.

## Conclusion

There is very low quality evidence to suggest that either chloroquine or hydroxychloroquine neither improves mortality or clinical course nor does it hasten virologic clearance in the treatment of COVID-19.

### Implications for research

Since the COVID-19 pandemic is ongoing, and >7.1 million patients have been infected worldwide, there is an urgent need to generate robust evidence regarding the efficacy and safety of CQ and HCQ in COVID-19. Randomized controlled studies of adequate sample size with sound methodology are needed to provide definite answers.

## Data Availability

The present study is a systematic review and meta-analysis

## References

1. Coronavirus Update (Live): 7,132,732 Cases and 406,959 Deaths from COVID-19 Virus Pandemic - Worldometer [Internet]. [cited 2020 Jun 8]. Available from: https://www.worldometers.info/coronavirus/?utm_campaign=homeAdUOA?Si,

2. RevisedNationalClinicalManagementGuidelineforCOVID1931032020.pdf [Internet]. [cited 2020 May 29]. Available from: https://www.mohfw.gov.in/pdf/RevisedNationalClinicalManagementGuidelineforCOVID1931032020.pdf

3. CDC. Coronavirus Disease 2019 (COVID-19) [Internet]. Centers for Disease Control and Prevention. 2020 [cited 2020 Apr 28]. Available from: https://www.cdc.gov/coronavirus/2019-ncov/hcp/therapeutic-options.html

4. Higgins JPT, Altman DG, Gøtzsche PC, Jüni P, Moher D, Oxman AD, et al. The Cochrane Collaboration’s tool for assessing risk of bias in randomised trials. BMJ. 2011 Oct 18;343:d5928.

5. Ottawa Hospital Research Institute [Internet]. [cited 2020 Jun 11]. Available from: http://www.ohri.ca/programs/clinical_epidemiology/oxford.asp

6. Guyatt GH, Oxman AD, Vist GE, Kunz R, Falck-Ytter Y, Alonso-Coello P, et al. GRADE: an emerging consensus on rating quality of evidence and strength of recommendations. BMJ. 2008 Apr 26;336(7650):924–6.

7. Borba MGS, Val F de A, Sampaio VS, Ara&amp MA, Alexandre U, Melo GC, et al. Chloroquine diphosphate in two different dosages as adjunctive therapy of hospitalized patients with severe respiratory syndrome in the context of coronavirus (SARS-CoV-2) infection: Preliminary safety results of a randomized, double-blinded, phase IIb clinical trial (CloroCovid-19 Study). medRxiv. 2020 Apr 16;2020.04.07.20056424.

8. Chen X, Zhang Y, Zhu B, Zeng J, Hong W, He X, et al. Associations of clinical characteristics and antiviral drugs with viral RNA clearance in patients with COVID-19 in Guangzhou, China: a retrospective cohort study. medRxiv. 2020 Apr 14;2020.04.09.20058941.

9. Mehra MR, Desai SS, Ruschitzka F, Patel AN. Hydroxychloroquine or chloroquine with or without a macrolide for treatment of COVID-19: a multinational registry analysis. The Lancet [Internet]. 2020 May 22 [cited 2020 May 23];0(0). Available from: https://www.thelancet.com/journals/lancet/article/PIIS0140-6736(20)31180-6/abstract

10. Kim MS, Jang S-W, Park Y-K, Kim B, Hwang T-H, Kang SH, et al. Treatment Response to Hydroxychloroquine, Lopinavir/Ritonavir, and Antibiotics for Moderate COVID 19: A First Report on the Pharmacological Outcomes from South Korea. medRxiv. 2020 May 18;2020.05.13.20094193.

11. Auld S, Caridi-Scheible M, Blum JM, Robichaux CJ, Kraft CS, Jacob JT, et al. ICU and ventilator mortality among critically ill adults with COVID-19. medRxiv. 2020 Apr 26;2020.04.23.20076737.

12. Yu B, Wang DW, Li C. Hydroxychloroquine application is associated with a decreased mortality in critically ill patients with COVID-19. medRxiv. 2020 May 1;2020.04.27.20073379.

13. Chen Jun LD, Chen Jun LD. A pilot study of hydroxychloroquine in treatment of patients with common coronavirus disease-19 (COVID-19). J Zhejiang Univ Med Sci. 2020 Mar 6;49(1):0–0.

14. Feng Z, Li J, Yao S, Yu Q, Zhou W, Mao X, et al. The Use of Adjuvant Therapy in Preventing Progression to Severe Pneumonia in Patients with Coronavirus Disease 2019: A Multicenter Data Analysis. medRxiv. 2020 Apr 10;2020.04.08.20057539.

15. Gautret P, Lagier J-C, Parola P, Hoang VT, Meddeb L, Mailhe M, et al. Hydroxychloroquine and azithromycin as a treatment of COVID-19: results of an open-label non-randomized clinical trial. Int J Antimicrob Agents. 2020 Mar 20;105949.

16. Geleris J, Sun Y, Platt J, Zucker J, Baldwin M, Hripcsak G, et al. Observational Study of Hydroxychloroquine in Hospitalized Patients with Covid-19. N Engl J Med. 2020 May 7;0(0):null.

17. Mallat J, Hamed F, Balkis M, Mohamed MA, Mooty M, Malik A, et al. Hydroxychloroquine is associated with slower viral clearance in clinical COVID-19 patients with mild to moderate disease: A retrospective study. medRxiv. 2020 May 2;2020.04.27.20082180.

18. Magagnoli J, Narendran S, Pereira F, Cummings T, Hardin JW, Sutton SS, et al. Outcomes of hydroxychloroquine usage in United States veterans hospitalized with Covid-19. medRxiv. 2020 Apr 23;2020.04.16.20065920.

19. Mahévas M, Tran V-T, Roumier M, Chabrol A, Paule R, Guillaud C, et al. Clinical efficacy of hydroxychloroquine in patients with covid-19 pneumonia who require oxygen: observational comparative study using routine care data. BMJ. 2020 May 14;369:m1844.

20. Huang M, Li M, Xiao F, Liang J, Pang P, Tang T, et al. Preliminary evidence from a multicenter prospective observational study of the safety and efficacy of chloroquine for the treatment of COVID-19. medRxiv. 2020 May 4;2020.04.26.20081059.

21. Rosenberg ES, Dufort EM, Udo T, Wilberschied LA, Kumar J, Tesoriero J, et al. Association of Treatment With Hydroxychloroquine or Azithromycin With In-Hospital Mortality in Patients With COVID-19 in New York State. JAMA [Internet]. 2020 May 11 [cited 2020 May 29]; Available from: https://jamanetwork.com/journals/jama/fullarticle/2766117

22. Tang W, Cao Z, Han M, Wang Z, Chen J, Sun W, et al. Hydroxychloroquine in patients with mainly mild to moderate coronavirus disease 2019: open label, randomised controlled trial. BMJ. 2020 14;369:m1849.

23. Chen Z, Hu J, Zhang Z, Jiang S, Han S, Yan D, et al. Efficacy of hydroxychloroquine in patients with COVID-19: results of a randomized clinical trial. medRxiv. 2020 Apr 10;2020.03.22.20040758.

24. Ip A, Berry DA, Hansen E, Goy AH, Pecora AL, Sinclaire BA, et al. Hydroxychloroquine and Tocilizumab Therapy in COVID-19 Patients - An Observational Study. medRxiv. 2020 May 25;2020.05.21.20109207.

25. Singh S, Khan A, Chowdhry M, Chatterjee A. Outcomes of Hydroxychloroquine Treatment Among Hospitalized COVID-19 Patients in the United States-Real-World Evidence From a Federated Electronic Medical Record Network. medRxiv. 2020 May 19;2020.05.12.20099028.

26. Novales FJM de, Ramírez-Olivencia G, Estébanez M, Dios B de, Herrero MD, Mata T, et al. Early Hydroxychloroquine Is Associated with an Increase of Survival in COVID-19 Patients: An Observational Study. 2020 May 5 [cited 2020 Jun 13]; Available from: https://www.preprints.org/manuscript/202005.0057/v1

27. Rosenberg ES, Dufort EM, Udo T, Wilberschied LA, Kumar J, Tesoriero J, et al. Association of Treatment With Hydroxychloroquine or Azithromycin With In-Hospital Mortality in Patients With COVID-19 in New York State. JAMA. 2020 May 11;

28. Touret F, de Lamballerie X. Of chloroquine and COVID-19. Antiviral Res. 2020 May 1;177:104762.

29. Antiviral Therapy | Coronavirus Disease COVID-19 [Internet]. COVID-19 Treatment Guidelines. [cited 2020 May 30]. Available from: https://www.covid19treatmentguidelines.nih.gov/antiviral-therapy/

30. Mehra MR, Ruschitzka F, Patel AN. Retraction—Hydroxychloroquine or chloroquine with or without a macrolide for treatment of COVID-19: a multinational registry analysis. The Lancet [Internet]. 2020 Jun 5 [cited 2020 Jun 8];0(0). Available from: https://www.thelancet.com/journals/lancet/article/PIIS0140-6736(20)31324-6/abstract

31. Million M, Gautret P, Colson P, Roussel Y, Dubourg G, Chabriere E, et al. Clinical Efficacy of Chloroquine derivatives in COVID-19 Infection: Comparative meta-analysis between the Big data and the real world. New Microbes New Infect. 2020 Jun 6;100709.

